# Life course longitudinal growth and risk of knee osteoarthritis at age 53 years: evidence from the 1946 British birth cohort study

**DOI:** 10.1101/2020.08.18.20177485

**Authors:** Katherine A. Staines, Rebecca Hardy, Hasmik J. Samvelyan, Kate A. Ward, Rachel Cooper

## Abstract

**Objectives:** To examine the relationship between height gain across childhood and adolescence with knee osteoarthritis in the MRC National Survey of Health and Development (NSHD).

**Methods:** Data are from 3035 male and female participants of the NSHD. Height was measured at ages 2, 4, 6, 7, 11 and 15 years, and self-reported at ages 20 and 26 years. Associations between (i) height at each age (ii) height gain during specific life periods (iii) Super-Imposition by Translation And Rotation (SITAR) growth curve variables of height size, tempo and velocity, and knee osteoarthritis at 53 years were tested.

**Results:** In sex-adjusted models, taller height at 4 and 6 years were modestly associated with decreased odds of knee osteoarthritis at age 53 (ORs per 1cm increase in height at age 6: 0.97 and age 4: 0.98 (95% CI: 0.95-1.00)). These associations were attenuated after adjustment for potential confounders. Similarly, taller adult achieved height measured at 26 and 53 years of age were associated with decreased odds of knee osteoarthritis (OR per 1cm increase in height: 0.98 (95% CI 0.96 to 1.00)). No associations were found between height gain during specific life periods or the SITAR growth curve variables and odds of knee osteoarthritis.

**Conclusions:** There was some limited evidence to suggest that taller height in childhood is associated with decreased odds of knee osteoarthritis at age 53 years in this cohort. This work enhances our understanding of osteoarthritis predisposition and the contribution of life course height to this.

## Introduction

Joint health is reliant upon the preservation of the articular cartilage and, its degradation is one of the main hallmarks of the degenerative joint disease osteoarthritis. Osteoarthritis, characterised by articular cartilage loss, subchondral bone thickening and osteophyte formation, is a major health care burden throughout the world. It is estimated that worldwide at least 10% of men and 18% of women aged over 60 years have symptomatic osteoarthritis. Osteoarthritis causes much pain and disability, and yet its underlying molecular mechanisms are not fully understood. Indeed, even the precipitating pathology remains a matter of debate and we are still unable to identify those at most risk of developing the disease.

Our previous work in a spontaneous murine model of ageing-related osteoarthritis, the STR/Ort mouse, revealed accelerated long bone growth, increased growth plate chondrocyte differentiation, and widespread abnormal expression of chondrocyte markers in osteoarthritis-prone mice.[1] Furthermore, we revealed enriched growth plate bridging, indicative of advanced and thus premature growth plate closure, in these mice.[1] Together this suggested that osteoarthritis development is associated with an accelerated growth phenotype and advanced pubertal onset.

Consistent with this finding, canine hip dysplasia (a hereditary predisposition to degenerative osteoarthritis) is more common in certain breeds, in particular larger breeds which tend to grow more rapidly.[2] However, associations between lifetime linear growth, i.e. height gain during specific life periods up to the attainment of adult height, and knee osteoarthritis development in human populations have, to our knowledge, not yet been studied. Previous epidemiological analyses of the Hertfordshire Cohort Study and the Medical Research Council National Survey of Health and Development (MRC NSHD) have found associations between low birth weight and high body mass index across life and increased risk of developing osteoarthritis.[3,4] This therefore suggests that life course size may predispose to osteoarthritis later in life.

Herein, we use one of these studies, the MRC NSHD, to examine the relationship between childhood and adolescent height growth and knee osteoarthritis at 53 years. Our aims were to: (1) test associations between height at different ages in early life and knee osteoarthritis in adulthood; (2) assess how patterns of height growth during childhood and adolescence are associated with knee osteoarthritis.

## Methods

### Study sample

The MRC NSHD is a birth cohort study, which includes a nationally representative sample of 2815 men and 2547 women born in England, Scotland, and Wales during 1 week in March 1946. The cohort has been followed prospectively across life with outcome data for these analyses drawn from a data collection in 1999, when participants were 53 years old.[5] At 53, 3035 participants (1472 men, 1563 women) participated, the majority (n = 2989) were interviewed and examined in their own homes by research nurses with others completing a postal questionnaire (n = 46). The responding sample at age 53 is in most respects representative of the national population of a similar age.[6] The data collection at age 53 years received ethical approval from the North Thames Multi-centre Research Ethics Committee, and written informed consent was given by all respondents.

### Outcome – knee osteoarthritis

During the home visit at age 53 years, trained nurses conducted clinical examinations of study participants’ knees.[3] Based on these examinations, the American College of Rheumatology criteria for the clinical diagnosis of idiopathic knee osteoarthritis were used to identify those with knee pain in either knee on most days for at least 1 month in the last year prior to the examination in 1999, and at least two of the following: stiffness, crepitus, bony tenderness and bony enlargement.[7]

### Height variables

Height was measured by nurses using standardised protocols at ages 2, 4, 7, 11 and 15 years, and self-reported at ages 20 and 26. Individual patterns of height growth during puberty were estimated using the SuperImposition by Translation and Rotation (SITAR) model of growth curve analysis, as previously described by Cole et al.[9,10] The SITAR model estimates the mean growth curve and three individual-specific parameters: size (reflecting differences in mean height), tempo (reflecting differences in the timing of the pubertal growth spurt) and velocity (reflecting differences in the duration of the growth spurt), each expressed relative to the mean curve.

### Covariates

Factors that may potentially confound the main associations of interest were selected *a priori* based on previous findings in the literature.[3] These were occupational class, sporting ability in adolescence, and leisure time physical activity at 53 years. As an indicator of socioeconomic position (SEP), occupational class at age 53 years (or if not available, the most recent measure in adulthood) was categorised into two groups – non manual (professional, managerial/ technical, skilled non-manual) and manual (skilled manual, partly skilled, unskilled) using the Registrar General’s Social Classification.[11] At age 13 years, children were graded as above average, average, or below average according to teacher reports of their sporting ability. This measure is used as a marker of motor skills and coordination evidenced by school-based physical activity and is predictive of leisure-time physical activity levels in adulthood.[12] Participation in leisure-time physical activity at age 53 was ascertained by asking participants to report whether they had undertaken any sports, vigorous leisure activities, or exercises in their spare time, not including getting to and from work, in the last 4 weeks prior to the examination and if so on how many occasions they had done these activities. This was categorised into three groups: inactive (no participation); moderately active (participated 1 to 4 times); most active (participated ≥5 times).

### Statistical analysis

To address the two main aims, we used logistic regression models to test associations between: (1) height at each age (aim 1); (2) conditional changes in height during specific life periods (early childhood: 2–4 years; late childhood: 4-7 years; childhood to adolescence: 7–15 years; adolescence to young adulthood: 15–20 years) (aim 2) and; (3) each SITAR height variable (aim 2) and odds ratios (ORs) of knee osteoarthritis. In initial models, we formally tested for interactions between sex and each main independent variable and where no evidence of interaction was found, models were fitted with men and women combined and adjustment for sex. In each set of models we first adjusted for sex (where there was no evidence of interaction) before then also adjusting for early and adult life factors (sporting ability + physical activity + occupational class). In models to address aim 2, we generated conditional changes in height by regressing each height measure on the earlier height measure for each sex and calculating the residuals.[13] The residuals were standardized (to have mean 0 and SD of 1) to ensure their comparability and these were included as the main independent variables. To maximise statistical power each set of models were run on the sample with valid data for the outcome, the specified independent variable and covariates. Data were analysed using Stata statistical software (version SE 14.2).

### Sensitivity analyses

In additional models we adjusted for adult weight at 53 years, considered a potential mediator. We also adjusted for weight at each age for aim 1, conditional weight gain (aim 2) and the SITAR weight variables (aim 2) to assess the contribution of weight during growth. To assess the potential impact of having to exclude those participants lost to follow-up and with missing data, comparisons were drawn between those included and those excluded in the main analyses. In addition, the sex-adjusted analyses were rerun in the maximum available samples including all available participants rather than being restricted to the sample with valid data on all measures. To assess the influence of potential secondary osteoarthritis on our findings the main analyses were repeated after excluding those participants with knee osteoarthritis who had reported ever seeing a doctor about an injury to the knee in which osteoarthritis was diagnosed.

## Results

### Cohort characteristics

A total of 1437 men and 1478 women had complete data on the SITAR parameters of height and knee osteoarthritis. Descriptive statistics are described in Table 1. In this sample, the percentage of individuals with knee osteoarthritis at 53 years of age was higher in women (13.04%) than in men (7.36%).

**Table 1:** Characteristics of the sample from the MRC National Survey of Health and Development with complete data on the SITAR parameters of height and the outcome, knee osteoarthritis. Knee osteoarthritis, social class and physical activity reported at 53 years, sporting ability at 13 years.

|  |  | Men (Max n=1437) |  |  | Women (Max n=1478) |  |  |
| --- | --- | --- | --- | --- | --- | --- | --- |
|  |  | n | Mean | SD | n | Mean | SD |
| Height 2 years (cm) |  | 1211 | 85.91 | 5.24 | 1197 | 84.72 | 4.57 |
| Height 4 years (cm) |  | 1288 | 103.51 | 5.10 | 1307 | 102.84 | 5.05 |
| Height 6 years (cm) |  | 1238 | 114.46 | 5.25 | 1255 | 113.74 | 5.26 |
| Height 7 years (cm) |  | 1249 | 120.35 | 5.65 | 1303 | 119.65 | 5.50 |
| Height 11 years (cm) |  | 1230 | 140.62 | 6.73 | 1257 | 141.16 | 6.94 |
| Height 15 years (cm) |  | 1135 | 162.04 | 8.86 | 1156 | 158.65 | 6.22 |
| Height 20 years (cm) |  | 1155 | 176.76 | 6.72 | 1231 | 162.62 | 6.24 |
| Height at 53 years (cm) |  | 1423 | 174.59 | 6.56 | 1467 | 161.58 | 5.97 |
|  |  | n | % |  | n | % |  |
| Knee osteoarthritis |  | 105 | 7.31 |  | 193 | 13.06 |  |
| Sporting ability: | Above average | 235 | 18.98 |  | 220 | 17.31 |  |
|  | Average | 793 | 64.05 |  | 902 | 70.97 |  |
|  | Below average | 210 | 16.96 |  | 149 | 11.72 |  |
| Physical activity: | Inactive | 693 | 48.26 |  | 748 | 50.61 |  |
|  | Moderately active | 268 | 18.66 |  | 240 | 16.24 |  |
|  | Most active | 475 | 33.08 |  | 490 | 33.15 |  |
| Social class: | Non-manual | 868 | 60.91 |  | 1044 | 71.21 |  |
|  | Manual | 557 | 39.09 |  | 422 | 28.79 |  |

### Life course height and knee osteoarthritis

In sex-adjusted models, taller height at age 6 (OR of knee osteoarthritis 0.97 per 1cm increase in height, 95% CI 0.95 to 1.00) was modestly associated with lower odds of knee osteoarthritis at age 53 years (Model 1; Table 2). There was also weak evidence of an association at age 4 (OR of knee osteoarthritis 0.98 per 1cm increase in height, 95% CI 0.95 to 1.00) (Table 2). However, with adjustment for early and adult life factors (Model 2), there was no longer clear evidence of associations at either of these ages (Table 2). Similarly, in sex-adjusted models taller adult achieved height measured at 53 years of age (or self-reported at 26 years) was associated with decreased odds of knee osteoarthritis (OR of knee osteoarthritis 0.98 per 1cm increase in height, 95% CI 0.96 to 1.00) (Table 2), however this too was attenuated when adjusted for early and adult life factors (Model 2). There was no evidence of association between height at any other age and knee osteoarthritis.

**Table 2:** Associations between height (per cm) at different ages throughout childhood, adolescence and young adulthood and odds ratios of knee osteoarthritis at age 53 years. Logistic regression Model 1: adjusted for sex; Model 2: further adjusted for sporting ability, physical activity and social class. There was little evidence of sex interaction in any of these models: P>0.09.

| Height (per cm) | n | Model | Odds ratio | 95% CI |  | p |
| --- | --- | --- | --- | --- | --- | --- |
| 2 years | 2085 | 1 | 0.99 | 0.96 | 1.02 | 0.5 |
|  |  | 2 | 1.00 | 0.97 | 1.03 | 0.8 |
| 4 years | 2281 | 1 | 0.98 | 0.95 | 1.00 | 0.07 |
|  |  | 2 | 0.99 | 0.96 | 1.02 | 0.4 |
| 6 years | 2232 | 1 | 0.97 | 0.95 | 1.00 | 0.05 |
|  |  | 2 | 0.99 | 0.96 | 1.01 | 0.3 |
| 7 years | 2321 | 1 | 0.98 | 0.96 | 1.01 | 0.1 |
|  |  | 2 | 0.99 | 0.97 | 1.02 | 0.6 |
| 11 years | 2314 | 1 | 0.99 | 0.97 | 1.01 | 0.5 |
|  |  | 2 | 1.00 | 0.98 | 1.02 | 0.9 |
| 15 years | 2150 | 1 | 0.99 | 0.97 | 1.01 | 0.5 |
|  |  | 2 | 1.00 | 0.98 | 1.02 | 1 |
| 20 years | 2144 | 1 | 0.99 | 0.96 | 1.01 | 0.2 |
|  |  | 2 | 0.99 | 0.97 | 1.02 | 0.6 |
| 26 years | 2231 | 1 | 0.98 | 0.96 | 1.00 | 0.08 |
|  |  | 2 | 0.99 | 0.97 | 1.01 | 0.3 |
| 53 years | 2474 | 1 | 0.98 | 0.96 | 1.00 | 0.03 |
|  |  | 2 | 0.99 | 0.97 | 1.01 | 0.3 |

### Height growth and knee osteoarthritis

No associations were found between height gains during any of the four periods assessed and odds of knee osteoarthritis at 53 years (Table 3). There was also no evidence of associations between height size, tempo or velocity (SITAR variables) and knee osteoarthritis at 53 years (Model 1; Table 4).

**Table 3:** Associations of conditional height gain (per standard deviation) during different periods of growth (early childhood: 2–4 years; late childhood: 4-7 years; childhood to adolescence: 7–15 years; adolescence to young adulthood: 15–20 years) with knee osteoarthritis at 53 years. Logistic regression Model 1: adjusted for sex; Model 2: further adjusted for sporting ability, physical activity and social class. There was little evidence of sex interaction in any of these models: P>0.06.

| Conditional change | n | Model | Odds ratio | 95% CI |  | p |
| --- | --- | --- | --- | --- | --- | --- |
| 2 - 4 years | 2004 | 1 | 0.91 | 0.78 | 1.05 | 0.2 |
|  |  | 2 | 0.96 | 0.83 | 1.12 | 0.6 |
| 4 - 7 years | 1885 | 1 | 0.93 | 0.80 | 1.08 | 0.3 |
|  |  | 2 | 0.96 | 0.83 | 1.11 | 0.6 |
| 7 - 15 years | 1862 | 1 | 1.10 | 0.94 | 1.27 | 0.2 |
|  |  | 2 | 1.10 | 0.95 | 1.28 | 0.2 |
| 15 - 20 years | 1752 | 1 | 1.03 | 0.88 | 1.20 | 0.7 |
|  |  | 2 | 1.04 | 0.89 | 1.22 | 0.6 |

**Table 4:** Associations between each parameter of the SITAR model of growth curve analysis (height size, tempo and velocity) and odds of knee osteoarthritis. Logistic regression Model 1: adjusted for sex Model 2: further adjusted for sporting ability, physical activity and social class. There was little evidence of sex interaction in any of these models: P>0.5

| SITAR variable (n=2493) | Model | Odds ratio | 95% CI |  | p |
| --- | --- | --- | --- | --- | --- |
| Size (cm) | 1 | 0.98 | 0.96 | 1.01 | 0.2 |
|  | 2 | 0.99 | 0.97 | 1.02 | 0.6 |
| Tempo (%) | 1 | 1.00 | 0.98 | 1.01 | 0.6 |
|  | 2 | 0.99 | 0.98 | 1.01 | 0.5 |
| Velocity (%) | 1 | 1.00 | 0.99 | 1.01 | 0.9 |
|  | 2 | 1.00 | 0.99 | 1.01 | 0.5 |

### Sensitivity analyses

Comparison of the characteristics of those individuals with complete data, vs those excluded are described in Tables S1.1 & S1.2. We found that higher proportions of those included were female (50.7% vs 43.7%; p< 0.001; Tables S1.1 & S1.2). No significant differences were observed in height between ages 2 – 15 years but at age 20, those included reported shorter heights than those excluded (171.0 cm vs 169.4 cm; Tables S1.1 & S1.2).

When models were adjusted for adult weight and weight during growth, there were no substantive differences in findings when examining associations between height at different ages throughout childhood, adolescence and young adulthood, and odds ratios of knee osteoarthritis at age 53 years (Tables S2.1 & S3.1). Similarly, there were no substantive differences in findings when investigating associations between conditional height gain during different periods of growth, and odds ratios of knee osteoarthritis at age 53 years (Tables S2.2 & S3.2). However, both these adjustments increased the association between SITAR height size and knee osteoarthritis (Tables S2.3 & 3.3). Moreover, adjusting for weight tempo also increased the association between height tempo and knee osteoarthritis at 53 years of age (Table S3.3).

When models were rerun on the maximum available samples including all available participants (Tables S43.1 – S4.3), there were no substantive differences in findings.

When we excluded those participants with potential secondary knee osteoarthritis from our analyses, there were no substantive differences in associations between height (Table S5.1) or conditional height gain (Table S5.2) and primary knee osteoarthritis at 53 years. A weak association was observed between SITAR height size when adjusted for sex (OR of knee osteoarthritis 0.97 per 1cm increase in height, 95% CI 0.95 to 1.00) (Model 1; Table S5.3). This was however attenuated when adjusted for early and adult life factors (Model 2; Table S5.3).

## Discussion

In this nationally representative British birth cohort study, associations between greater height at ages 4 and 6 years and marginally lower odds knee osteoarthritis at age 53 were observed in sex-adjusted models, but these were attenuated after adjustment for early and adult life factors. No associations were observed between height changes during early childhood, late childhood, childhood to adolescence or adolescence to young adulthood or SITAR parameters and knee osteoarthritis.

A major strength of our study is the availability of multiple prospectively ascertained measurements of height throughout childhood and adolescence in the NSHD, together with the already derived SITAR variables and measures of knee osteoarthritis in a relatively large sample of people in midlife.[9] This provided a unique opportunity to investigate the associations between life course longitudinal growth and knee osteoarthritis. We used two approaches to model growth and understand its relation to knee osteoarthritis in later life. Firstly, we used a conditional change approach to enable us to determine whether there are specific sensitive period/s of growth which may be associated with knee osteoarthritis. This can be interpreted as the change in height size above or below that expected given earlier height, and thus is useful in identifying accelerated or restricted growth.[14] We next chose the SITAR growth curve model since it was previously shown to effectively summarise pubertal growth based on three parameters of size, velocity and tempo.[9,10]

Our previous work explored associations between growth dynamics and osteoarthritis onset in a spontaneous murine model of osteoarthritis, the STR/Ort mouse.[1] We revealed accelerated long bone growth, aberrant expression of growth plate markers and enriched growth plate bridging, indicative of advanced and thus premature growth cessation, in these osteoarthritis-prone mice.[1] Together this suggested that these accelerated growth dynamics in young osteoarthritis-prone mice may underpin their osteoarthritis onset. However, whether these observations are unique to osteoarthritis in the STR/Ort mouse or are characteristic of human osteoarthritis in general had yet to be established. This study suggests that in the NSHD, associations between greater gains in height, indicative of accelerated growth, are not associated with increased odds of knee osteoarthritis. Rather, the modest associations found suggest the opposite. It is however important to note that this was examined in midlife when the cohort are still relatively young, and osteoarthritis prevalence (7.31% in men; 13.06% in women) is lower than that seen currently in primary care at this age. It would therefore be of interest to further examine these potential associations in older individuals.

Primary osteoarthritis is described as naturally occurring or ageing-related osteoarthritis, while secondary osteoarthritis is associated with other causes including trauma. Our previous findings in the STR/Ort mouse examined primary murine osteoarthritis [1] and therefore to examine the influence of secondary knee osteoarthritis on the patterns of height growth in the NSHD, we ran a sensitivity analysis in which we excluded individuals who had reported consulting a Doctor about a knee injury. However, whilst we found no substantive differences in findings, this highlights the need to examine the risk of osteoarthritis in aged individuals where primary knee osteoarthritis is more prevalent.

Our study extends a previous study examining this British birth cohort in which prolonged exposure to high BMI through adulthood increased risk of development of knee osteoarthritis at age 53.[3] This is consistent with our sensitivity analyses in which adjustment for weight strengthened the associations between SITAR height size and odds of knee osteoarthritis. Wills et al., also found that BMI increases from childhood to adolescence (7–15 years) were positively associated with knee osteoarthritis, however this was in women only.[3] In our analyses, we found no evidence of differences in association by sex. We did find that in our cohort with complete data, women had a higher prevalence of knee osteoarthritis, similar to that reported previously in the NSHD, and in primary care.[3,15] Wills et al., concluded that the excessive weight during this period may result in altered mechanical loading to the knee joint. Similarly, it is likely that periods of accelerated growth will also impact on the biomechanics of the joint. The shape of the hip joint is largely determined in childhood, and previous studies have identified that in the NSHD, this is associated with (i) age of onset of walking in infancy [16] (ii) higher BMI at all ages and greater gains in BMI [17] and (iii) height, weight, BMI and BMD at ages 60-64 years.[18] Similarly, in the Avon Longitudinal Study of Parents and Children (ALSPAC) cohort, hip shape in perimenopausal women is associated with hip osteoarthritis susceptibility loci and may contribute to hip osteoarthritis later in life.[19] Recent evidence in the ALSPAC cohort has also identified pubertal timing, as reflected by height tempo, to be associated with hip shape.[20] Further, in the UK Biobank, early menarche is associated with higher risk for osteoarthritis.[21] However these associations were not observed in this study.

Together, in this relatively large population-based cohort study, there was limited evidence to suggest that height in childhood is associated with odds of knee osteoarthritis at age 53 years. However, there were no associations with height gain during specific periods of growth, or with the SITAR height growth variables. This work enhances our understanding of osteoarthritis predisposition and the contribution of life course height to this.

## Data Availability

Data used in this publication are available to bona fide researchers upon request to the NSHD Data Sharing Committee via a standard application procedure. Further details can be found at http://www.nshd.mrc.ac.uk/data. doi: 10.5522/NSHD/Q101

## Acknowledgements & affiliations

The authors thank all the participants of the MRC National Survey of Health and Development and all staff involved in data collection and data entry. The authors would like to acknowledge the Medical Research Council for funding to KS (MR/R022240/1). The authors would also like to thank Dr Alex Ireland (Manchester Metropolitan University, UK) for his insightful discussions during the preparation of this manuscript.

